# Questionnaire assessment helps the self-management of patients with inflammatory bowel disease during the outbreak of Coronavirus Disease 2019

**DOI:** 10.1101/2020.03.25.20043364

**Authors:** Meiping Yu, Zhenghao Ye, Yu Chen, Tingting Qin, Jiguang Kou, De’an Tian, Fang Xiao

**Affiliations:** Department of Gastroenterology, Tongji Hospital, Tongji Medical College, Huazhong University of Science and Technology, Wuhan, China; Department of Biliary – Pancreatic Surgery, Tongji Hospital, Tongji Medical College, Huazhong University of Science and Technology, Wuhan, China; Department of Gastroenterology, Xiaogan Hospital Affiliated to Wuhan University of Science and Technology, Wuhan, China

**Keywords:** questionnaire, inflammatory bowel disease, Coronavirus Disease 2019

## Abstract

**Background and Aims:** The outbreak of Coronavirus Disease 2019 (COVID-19) may affect the disease status of patients with inflammatory bowel disease (IBD). This study aimed to assess the disease status of IBD patients in Hubei province by questionnaire online and guide to the self-management of IBD patients during this epidemic.

**Methods:** A questionnaire was designed containing the Harvey-Bradshaw Index (HBI), the Partial Mayo Score (PMS), the short inflammatory bowel disease questionnaire (SIBDQ) and distributed to Hubei IBD patients online within one month of traffic control after the outbreak of COVID-19. This questionnaire also included some questions about patients’ self-report disease conditions and their epidemiological history of COVID-19.

**Results:** A total of 102 eligible questionnaires were included in the analysis. No patient reported infection with severe acute respiratory syndrome coronavirus 2 (SARS-CoV-2) in our study. Our result showed that 69.64% of patients with ulcerative colitis (UC) and 80.44% of patients with Crohn’s disease (CD) were in remission. There was not a statistically significant difference in the proportion of the active disease stage between the two types of disease (p=0.103). The majority of patients (85.29%) had a good health-related quality of life (HRQoL) (SIBDQ≥50). The reduction in physical exercise is a risk factor for worsening in conditions (OR=17.593, 95%CI 2.035 to 152.097, p=0.009).

**Conclusions:** The outbreak of COVID-19 might not have a significant impact on most Hubei IBD patients within one month after the traffic control. The patient’s disease condition could be assessed by our questionnaires. Doctors utilized the information and advised for IBD patients about self-management during the period of COVID-19.

## 1. Introduction

In December 2019, an outbreak of Coronavirus Disease 2019 (COVID-19) caused by the SARS-COV-2 emerged in Wuhan, the capital of Hubei Province, China, and spread rapidly worldwide^1,2^. On January 23, 2020, the Chinese government implemented traffic controls to prevent the spread of COVID-19^3^. Up to 20 February 2020 approximately a month after traffic control when we distributed this questionnaire to IBD patients, there had been 75,465 cases of COVID-19 confirmed in mainland China, including 62,662 cases in Hubei Province. The number of confirmed diagnoses in Hubei Province is 83.03% of that in China, accounting for the vast majority. At the same time, medical resources had also shifted more towards the diagnosis and treatment of COVID-19 in Hubei province. These factors made routine medical treatment and follow-up of patients with chronic diseases inconvenient. Under these influences, it is essential for patients with chronic diseases to self manage under the guidance of a doctor in this particular period. Self-management is the process by which patients participate in decision-making and self-care under the guidance of a doctor^4^. Patient’s effective self-management can relieve symptoms and control disease activity to a certain extent^5^.

Inflammatory bowel disease (IBD) is a type of chronic idiopathic bowel disease, includes Crohn’s disease (CD) and ulcerative colitis (UC). IBD patients have varying degrees of immune disorders^6^ so that they may be considered as virus-susceptible. It is particularly crucial for IBD patients to know how to self manage during the epidemic. Self-management requires monitoring of diseases by doctors first^7^. Our questionnaire can assess patients’ disease activity and quality of life online, which is an effective and convenient way to guide patients to promote better self-management.

The questionnaire was a validated 60-item questionnaire including scoring systems for PMS, HBI and SIBDQ. These indexes were designed to gain detailed knowledge of disease activity and the quality of life of these IBD patients. There were questions to understand subjective feelings of disease conditions. This questionnaire also included questions about COVID-19 epidemiological history of these IBD patients. After evaluating the patient’s disease activity and HRQoL by those indexes, we gave feedback to these IBD patients and guided patients to develop targeted self-management programs. This approach might facilitate the development and modification of interventions to optimize patient self-management during the COVID-19 epidemic^8^.

## 2. Materials and Methods

### 2.1. Study Design and Participants

This study was an online questionnaire survey among IBD patients from the region of Hubei province. From February 18, 2020 to February 20, 2020, the questionnaire was administered to a sample of IBD patients with regular follow-up in our IBD center. There were two items of the inclusion criteria for this study. One was that the patient (aging from 14 to 80) was established diagnosis of IBD for at least three months, another was that the patient is available to finish the online questionnaire (by Wechat, QQ, website, email) by himself or with the help of others. The exclusion criterion was that the patient is not able to finish the questionnaire. During this study, patients with IBD had been informed of the study’s aim from their GE. An IBD specialist nurse is explicitly trained in this questionnaire contacted them to explain the study objectives. Participants completed the questionnaire with an online survey portal. They completed questionnaires voluntarily and independently, under uncompensated conditions.

### 2.2. Questionnaire design

The questionnaire was a validated 60-item questionnaire that assesses IBD disease activity across multiple domains, including the HBI, PMS and SIBDQ. HBI score ranges from 0 to 16 or more, and the highest score depends on the number of liquid stools per day. HBI scores of 0 and 4 are assigned to the remission phase; 5 to 8 are assigned to moderately active disease phase; ≥9 are assigned to severely active disease phase^9^. We used PMS (excludes the endoscopic appearance of the mucosa), a 9-point score composed of the stool frequency, bleeding components and physician rating of disease activity. PMS of 0 and 2 are assigned to the remission phase; 3 to 6 are assigned to moderately active disease phase; 7 to 9 are assigned to severely active disease phase^10^. Total SIBDQ score ranges from 10 to 70, to find out how the patients have been feeling during the last two weeks. They will be asked about symptoms related to IBD diagnosis, as well as the general emotion status during the period and their attitude to the plague. Patients with a SIBDQ score of more than 50 are considered to have a good HRQoL^11^.

In addition to the question about HBI, PMS and SIBDQ, the questionnaire included questions regarding the subjective feeling about their change in disease conditions and epidemiological history questions about COVID-19.

### 2.3. Statistical analysis

Data collection and its statistical analysis were carried out using the SPSS software system (SPSS for Windows, Version 23.0, SPSS Inc., Chicago). Data analysis excluded incomplete items. Categorical data obtained were presented as frequency counts and percentages. Median, range and frequency were used to describe the demographic, SIBDQ scores and clinical characteristics. Chi-square tests were used to test the significant difference of categorical variables between two groups, such as the participants’ distribution in different disease active phase between UC and CD patients, with fisher exact test as appropriate. The logistic regression analysis was used to identify variables significantly associated with disease activity, and a multivariate model was built to assess the effect of each potential confounding factor and determined independent and significant factors associated with the disease index. The odds ratio (OR) was calculated to quantify the corresponding risk. For all analyses, p<0.05 was considered statistically significant.

### 2.4. Ethical Considerations

This study was approved by the National Health Commission of China and Ethics Commission of Tongji Hospital, Tongji Medical College, Huazhong University of Science and Technology.

## 3. Results

### 3.1. Presenting Characteristics

A total of 111 electronic questionnaires were returned. There were 102 valid questionnaires, with an effective rate of 91.89%. The nine questionnaires excluded were due to some missing items. The median age of participants was 34 years (IQR, 27.25-42.25; range, 14-66), and 66.67% of participants were men. There were 56 (54.90%) patients with ulcerative colitis and 46 (45.10%) patients with Crohn’s disease. Among all the participants in our survey, no one reported infection with SARS-CoV-2; no one had symptoms related to COVID-19 or had a history of exposure. Table 1 shows the demographic data and disease-related variables for all participants who agreed and completed the survey.

**Table 1.** Baseline characteristics of the study population. (n=102).

| Characteristics | Value |
| --- | --- |
| Age, Median (min-max) (IQR), y | 34 (14-66) (27.25-42.25) |
| Gender |  |
| Female | 34 (33.33%) |
| Male | 68 (66.67%) |
| Diagnosis |  |
| Ulcerative Colitis | 56 (54.90%) |
| Crohn's Disease | 46 (45.10%) |
| Diagnosed with COVID-19 | 0 |
| Huanan seafood market exposure | 0 |
| Signs and symptoms of COVID-19 | 0 |
| Habitation* |  |
| Wuhan | 26 (25.49%) |
| Xiaogan | 22 (21.57%) |
| Jingzhou | 12 (11.76%) |
| Suizhou | 8 (7.83%) |
| Xiangyang | 6 (5.88%) |
| Huangshi | 6 (5.88%) |
| Huanggang | 5 (4.90%) |
| Yichang | 5 (4.90%) |
| Jingmen | 4 (3.92%) |
| Xianning | 2 (1.96%) |
| Xiantao | 2 (1.96%) |
| Enshi Tujia and Miao Autonomous Prefecture | 2 (1.96%) |
| Tianmen | 1 (0.98%) |
| Qianjinag | 1 (0.98%) |

\*.All participants are located in Hubei Province

### 3.2. Disease activity

We used PMS and HBI to score and grade disease activity in UC and CD patients, respectively. The results were shown in Table 2. The median PMS of UC patients was 2 (IQR, 0-3; range, 0-8). Of the 56 UC patients, 39 (69.64%) UC patients were in remission, 16 (28.57%) patients had moderate activity, and 1 (1.79%) patients had severe activity. The median HBI of CD patients was 2 (IQR, 1-4; range, 0-12). Of the 46 CD patients, 37 (80.44%) CD patients were in remission, 6 (13.04%) patients had moderate activity, and 3 (6.52%) patients had severe activity. There was not a statistically significant difference in the proportion of the disease activity stage between UC and CD patients (p = 0.103).

**Table 2.**
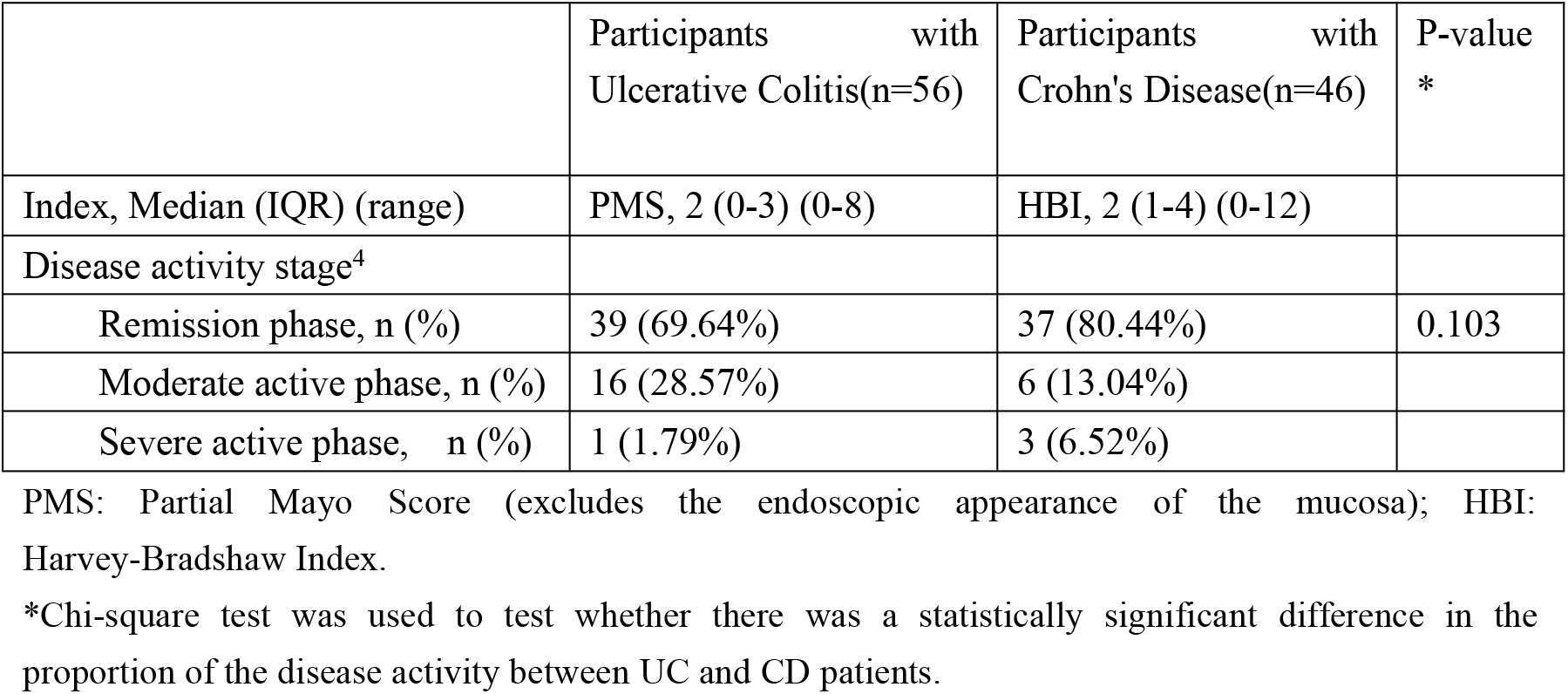
Evaluation of participants’ IBD disease activity

|  | Participants with<br>Ulcerative Colitis(n=56) | Participants with<br>Crohn's Disease(n=46) | P-value<br>* |
| --- | --- | --- | --- |
| Index, Median (IQR) (range) | PMS, 2 (0-3) (0-8) | HBI, 2 (1-4) (0-12) |  |
| Disease activity stage<sup>4</sup> |  |  |  |
| Remission phase, n (%) | 39 (69.64%) | 37 (80.44%) | 0.103 |
| Moderate active phase, n (%) | 16 (28.57%) | 6 (13.04%) |  |
| Severe active phase, n (%) | 1 (1.79%) | 3 (6.52%) |  |

PMS: Partial Mayo Score (excludes the endoscopic appearance of the mucosa); HBI: Harvey-Bradshaw Index.
\*Chi-square test was used to test whether there was a statistically significant difference in the proportion of the disease activity between UC and CD patients.

### 3.3. Quality of life

The median SIBDQ of all participants was 59 (IQR, 52.25-63; range, 34-70). The median SIBDQ of UC patients was 60 (IQR, 54.75-64; range, 35-70), and the median SIBDQ of CD patients was 58 (IQR, 52-62.75; range, 34-69). Among all participants, 87 (85.29%) patients had good HRQoL (SIBDQ ≥ 50). There were 49 (87.50%) UC patients who had good HRQoL, compared with 38 (82.61%) CD patients who had good HRQoL (p=0.338) (table 3), suggesting HRQoL is not significantly different between UC and CD patients.

**Table 3.** Evaluation of participants’ SIBDQ

|  | Total participants<br>(n=102) | Participants with UC and CD respectively |  |  |
| --- | --- | --- | --- | --- |
|  |  | Participants with<br>UC (n=56) | Participants with CD<br>(n=46) | P-value* |
| SIBDQ, Median (IQR) | 59 (52.25-63) | 60 (54.75-64) | 58 (52-62.75) |  |
| HRQoL |  |  |  | 0.338 |
| Good ($\geq 50$), n (%) | 87 (85.29%) | 49 (87.50%) | 38 (82.61%) | |
| Poor ($< 50$), n (%) | 15 (14.71%) | 7 (15.50%) | 8 (17.39%) | |

HRQoL: health-related quality of life
\*Chi-square test was used to test whether there was a statistically significant difference in the proportion of HRQoL between UC and CD patients.

### 3.4. Self-report disease conditions

In this questionnaire, we set up questions that allow patients to self-report changes in their disease condition. Approximately half of the patients (n = 55, 53.92%) thought that their disease condition did not change during the epidemic, 25 (24.51%) considered their disease condition improved, and 22 (21.57%) considered their disease condition worsening. We attempted to study the cause of change in patients’ self-report disease conditions. The result showed that reduced physical exercise is a risk factor for worse in the disease condition (OR=17.593, 95%CI 2.035 to 152.097, p=0.009). The other factors did not have a significant risk for changes in the patient’s disease condition. These data are shown in Table 4.

**Table 4.** Multi-variable logistic regression of the causes of change in participants’ self-evaluation disease condition.

| Disease condition | Characters | n (%) | OR (95%CI) | P-value |
| --- | --- | --- | --- | --- |
| Condition worsening<br>(n=22) | Subsequent visit |  |  |  |
|  | No | 18 (81.82%) | 3.785(0.871, 16.458) | 0.076 |
|  | Yes | 4 (18.18%) |  |  |
|  | Medication regimen |  |  |  |
|  | Not changed | 14 (63.64%) | 0.264 (0.060, 1.167) | 0.079 |
|  | Changed | 8 (36.36%) |  |  |
|  | Emotional status |  |  |  |
|  | Negative | 4 (18.18%) | 3.306(0.413, 26.454) | 0.260 |
|  | Positive | 3 (13.64%) | 0.830 (0.178, 3.880) | 0.813 |
|  | Normal | 15 (68.18%) |  |  |
|  | Physical exercise |  |  |  |
|  | Reduced | 5 (22.73%) | 17.593 (2.035, 152.097) | 0.009 |
|  | Not reduced | 17 (77.27%) |  |  |
|  | Rest |  |  |  |
|  | Inadequate | 21 (95.45%) | 1.071 (0.262, 4.378) | 0.924 |
|  | Adequate | 1 (4.55%) |  |  |
|  | Smoking |  |  |  |
|  | No | 19 (86.36%) | 0.665 (0.123, 3.591) | 0.636 |
|  | Yes | 3 (13.64%) |  |  |
| Condition improved<br>(n=25) | Subsequent visit |  |  |  |
|  | No | 21 (84.00%) | 2.730 (0.731, 10.199) | 0.135 |
|  | Yes | 4 (16.00%) |  |  |
|  | Medication regimen |  |  |  |
|  | Not changed | 19 (76.00%) | 0.374 (0.094, 1.496) | 0.165 |
|  | Changed | 6 (24.00%) |  |  |
|  | Emotional status |  |  |  |
|  | Negative | 1 (4.00%) | 2.094 (0.138, 31.710) | 0.594 |
|  | Positive | 14 (56.00%) | 2.259 (0.751, 6.792) | 0.147 |
|  | Normal | 10 (40.00%) |  |  |
|  | Physical exercise |  |  |  |
|  | Reduced | 1 (4.00%) | 0.694 (0.222, 2.167) | 0.529 |
|  | Not reduced | 24 (96.00%) |  |  |
|  | Rest |  |  |  |
|  | Inadequate | 9 (36.00%) | 0.136 (0.015, 1.249) | 0.078 |
|  | Adequate | 16 (64.00%) |  |  |
|  | Smoking |  |  |  |
|  | Yes | 4 (15.38%) | 0.481 (0.104, 2.228) | 0.350 |
|  | No | 22 (84.62%) |  |  |

\*The reference categories: Condition no change.

### 3.5. The change in medication regimens

We studied participants’ medication regimens before and after the outbreak of COVID-19. The results are shown in Table 5. Twenty patients who changed their medication regimens, with 10 UC patients and 10 CD patients. Then we investigated the reasons for the change in patient’s medication regimens during this outbreak situation. The largest proportion of the reason was the inability to purchase medications (n = 18, 90.00%), and the second reason was the inability to go to the hospital for routine treatment (n = 2, 10.00%). No one chose the options of “forgetting to take medicine” and “reducing medicine on your own”, which suggest that medication compliance of these IBD patients we surveyed was excellent during the epidemic.

**Table 5.** The comparison of the medication regimens before and after the outbreak of COVID-19 and the changes in medication regimens.

|  | Medication | Before the outbreak, n (%) | After the outbreak, n (%) | The number of changes*, n |
| --- | --- | --- | --- | --- |
| UC patients, (n=56) | No medication | 1 (1.75%) | 4 (5.26%) | +3 |
|  | Mesalazine | 50 (89.47%) | 48 (85.96%) | -2 |
|  | Remicade | 3 (5.26%) | 3 (5.26%) | 0 |
|  | Enteral nutrition | 1 (1.75%) | 1 (1.75%) | 0 |
|  | Glucocorticoid | 2 (3.51%) | 2 (3.51%) | 0 |
|  | Adalimumab | 1 (1.75%) | 1 (1.75%) | 0 |
|  | Probiotics | 4 (5.26%) | 4 (5.26%) | 0 |
| CD patients, (n=46) | No medication | 1 (1.92%) | 3 (5.77%) | +2 |
|  | Adalimumab | 1 (1.92%) | 2 (3.85%) | +1 |
|  | Remicade | 21 (46.15%) | 16 (36.54%) | -5 |
|  | Methotrexate | 2 (3.85%) | 1 (1.92%) | -1 |
|  | Mesalazine | 8 (17.31%) | 8 (17.31%) | 0 |
|  | Enteral nutrition | 10 (19.23%) | 10 (21.15%) | 0 |
|  | Azathioprine | 20 (40.38%) | 20 (40.38%) | 0 |
|  | Glucocorticoid | 2 (3.85%) | 2 (3.85%) | 0 |
|  | Thalidomide | 1 (1.92%) | 1 (1.92%) | 0 |
|  | Probiotics | 1 (1.92%) | 1 (1.92%) | 0 |

\*Plus sign indicates an increase in quantity and minus sign indicates a decrease in quantity.

### 3.6. Emotional state

Negative emotions are significantly correlated with clinical recurrence and are also considered to be independent risk factors for more frequent activities of disease^12,13^. Therefore, we also investigated the emotional states of IBD patients in this survey. More than half of the participants (57.85%) thought they had the ordinary moods during this epidemic, 35.29% had positive moods, and 6.86% had negative moods.

## 4. Discussion

COVID-19 broke out in December 2019 in China. Hubei Province is one of the worst-hit areas. COVID-19 was caused by SARS-CoV-2 which was generally susceptible to population^14^. The elderly and patients with underlying diseases were more susceptible to infection and prone to serious consequences^15^. Our results showed that there was no IBD patient reported being diagnosed with COVID-19 in this survey. But this did not mean that IBD patients were not susceptible to SARS-CoV-2 since the samples of this study were not obtained by random sampling and the sample size was relatively small. Until now, there is no evidence to prove the susceptibility of IBD patients to COVID-19 from other study^16^. The epidemic led to a contraction in routine medical resources, and IBD patients had difficulty maintaining previous disease management. Our research focuses on guiding patients’ self-management through the results of questionnaires.

Since the start of traffic control in China in late January, the lifestyle, psychological and physical condition of the Chinese population might change^17-19^, especially for residents in Hubei province. This situation made it more difficult for IBD patients in Hubei province to have face-to-face medical interventions. We used web questionnaires to evaluate the disease activity, quality of life, and the patient’s self-report disease condition of IBD patients. Then we provided feedback on the patient’s condition and advised on their self-management. This manner helped IBD patients to adjust their treatment plans and develop a home-based self-management medical intervention model. It has been proven that active doctor-patient communication can improve patient confidence with treatment, shared decision-making capacity and then has a good impact on disease activity^4,20^. We will continue to distribute questionnaires to this group of IBD patients in Hubei province every month until the policy of traffic control removed. Then we will compare the patient’s objective evaluation results with the subjective report results. The final goal is to guide patient home-based self-management during the outbreak of COVID-19.

Our results showed that 69.64% of UC patients and 80.44% of CD patients were in remission assessed by PMS and HBI index. 85.29% of patients had a good quality of life during the epidemic through the SIBDQ test. Among the patients’ self-reported results, 78.43% of the patients thought that their disease conditions had not changed or even improved. The disease activity and quality of life of most patients were still in an ideal state, and their self-reported disease conditions did not worsen. It showed that the epidemic had not a significant negative impact on the majority of IBD patients within one month of traffic control.

We studied the influencing factors of the self-report disease condition of IBD patients during this epidemic. The factors included “emotional state,” “change of medication regimen,” “daily rest,” “subsequent visit,” “reduction of exercise” and “smoking.” The results showed that reduced exercise is a risk factor for worsening disease (OR=17.593, p=0.009). The other factors that could affect the disease conditions of IBD patients in previous studies^21-26^, but have not shown a statistical correlation in our survey with a relatively small sample size. Therefore, we still recommend that IBD patients maintain a positive emotional state, maintain the medication regimen, have adequate rest, make timely doctor-patient communication, take proper-intensity exercise and quit smoking during the self-management process.

Pharmacological intervention is a key part of managing symptoms and maintaining remission in patients with IBD^27^. It had become inconvenient for patients to return to the hospital or take appropriate medications under this special circumstance. Our results showed that the number of people who took no medication increased among UC and CD patients after the outbreak of COVID-19. For UC patients, the number of patients taking mesalazine decreased due to the inconvenience of obtaining medications. For CD patients, the use of Remicade and methotrexate decreased, and the use of adalimumab increased. The decrease in medications might be due to the inconvenience of obtaining medications. We recommend using other available medicines instead of inaccessible Remicade. We tried to develop personalized treatment plans for specific patients on time in this particular period. Such a measure could minimize the impact of changes in medication regimens on the patient’s disease condition. It was an important step to increase patient self-management.

The result showed that 93.14% of patients had normal or even positive emotional states; only 6.86% of patients had negative emotional states in our survey. The result showed that the epidemic did not have a very negative impact on the mood of these IBD patients in such a relatively early month. However, previous studies have shown that emotional stress is significantly associated with decreased quality of life, interventions to address psychological issues may be part of IBD management^28,29^. Therefore, we should take intervention in those patients with negative moods to prevent the emotional states from adversely affecting their disease conditions.

## 5. Conclusion

In the middle of February, more than half of the Hubei IBD patients in our study were in remission. The majority of participates had a good HRQoL, and their self-reported disease condition did not worsen. This result indicated that the outbreak of COVID-19 might not have a significant impact on most IBD patients within one month after the traffic control. We will continue to pay attention to the long-term effect of the epidemic on disease activity and HRQoL in IBD patients in the future follow-up. In our survey, doctors used questionnaires to assess patients’ disease, give timely feedback and suggestions to IBD patients. This method facilitated the effective self-management of patients under the circumstance of the COVID-19 outbreak.

## Data Availability

All data, models, and code generated or used during the study appear in the submitted article.The data used to support the findings of this study are available from the corresponding author upon request.

## Abbreviations

COVID-19: Coronavirus Disease 2019
IBD: inflammatory bowel disease
HBI: Harvey-Bradshaw Index
PMS: Partial Mayo Score
SIBDQ: short inflammatory bowel disease questionnaire
SARS-COV-2: severe acute respiratory syndrome coronavirus 2
UC: ulcerative colitis
CD: Crohn’s disease
HRQoL: health-related quality of life
OR: odds ratio
IQR: interquartile range

## Acknowledgments

We thank all those who gave their time and efforts to participants in this study.

## Authors’ contributions

Meiping Yu designed the questionnaire; Meiping Yu, Zhenghao Ye, Yu Chen analysed data and wrote the manuscript; Tingting Qin guided the statistical method; Jiguang Kou contributed to the collection of samples; De’an Tian and Fang Xiao supervised the study; Fang Xiao designed the study concept and critically revised the manuscript; all authors reviewed and accepted the final version of the manuscript.

